# Post-stroke delirium and associations with neuroimaging biomarkers on routine Computed Tomography (CT): a cohort study

**DOI:** 10.1101/2024.05.28.24308083

**Authors:** Amanda Barugh, Andrew Farrall, Karen Ferguson, Susan Shenkin, Alasdair MacLullich, Gillian Mead

**Author notes:** Address correspondence to Professor Gillian Mead.

## Abstract

**Background:** Delirium affects a quarter of patients after acute stroke and predicts poorer outcomes. Our aim was to determine whether either qualitative assessment or quantitative assessments of the regional atrophy obtained from routinely performed computed tomography (CT) brain imaging could identify those most at risk of developing delirium.

**Methods:** We recruited 95 patients with acute stroke (age ≥65) over one year. Follow-up for delirium and cognition was performed at 1, 3, 5, 7, 14, 21, 28 days, 4 months and 12 months. All participants underwent routine CT brain (Toshiba 64-slice or 128-slice scanner). White matter disease and atrophy were rated qualitatively (mild, moderate, severe). Atrophy in multiple specific areas was measured quantitatively.

**Results:** Twenty-six (27%) developed delirium during the 12 months of follow-up. On univariable analysis, delirium was associated with increasing age, being female, less independent in pre-stroke activities of daily living, pre-existing cognitive impairment, increasing stroke severity, having had a total anterior circulation stroke and global cerebral atrophy on brain CT. Multivariable analysis demonstrated that only global cerebral atrophy, being female and having a more severe stroke predicted delirium. This model accounted for between 38% and 55% of the variance in delirium.

For quantitative CT analysis, on univariable analysis, delirium was associated with atrophy in several specific brain areas. On multivariable analysis, only NIHSS (for every one point increase OR 1.23, 95% CI 1.06-1.43; p=0.006)) and cistern ambiens ratio (OR 1.41, 95% CI 1.48-4.96; p=0.028) were significantly associated. This model accounted for between 35.1% and 51.2% of delirium variance.

**Conclusion:** Clinical variables together with either qualitative atrophy assessment or cistern ambiens ratio on routine CT brain could identify stroke patients most at risk of delirium and to stratify patients in clinical trials of delirium prevention and treatment.

## Background

Delirium is a serious, acute neuropsychological disorder [1] affecting around a quarter of patients with acute stroke [2–5]. It is associated with serious adverse outcomes including increased mortality, prolonged hospital stay and poorer long-term cognition [6]. Understanding how to improve cognition after stroke is priority for further research [7].

Delirium after stroke is associated with increasing age and stroke severity [4,8]. Most patients with acute stroke will have a plain CT scan to identify intracerebral haemorrhage and signs of ischaemia and old stroke lesions. CT can also identify white matter lesions (WMLs) and cerebral atrophy with very good agreement with magnetic resonance imaging [9,10].

A 2022 systematic review showed that delirium was associated with cortical lesions, anterior circulation, supratentorial, larger (versus small volume), cardioembolic aetiology and large vessel disease, but this review did not report associations with ‘background’ disease including old stroke lesions and background changes [11].

Other studies have sought associations between background atrophy (using qualitative measures of atrophy) and WMD but results vary [6,8,12,13]. To our knowledge no previous studies of delirium after stroke have measured atrophy in pre-defined areas of the brain (either using callipers for hard copies of scans or computer graphical functions for electronic scans). In intensive care survivors, atrophy in specific brain regions was associated with delirium duration [14]. Quantitative linear measurements of atrophy on CT have been used to detect (n=129) [15] and diagnose (n=307) [16] Alzheimer’s Disease (AD).

Quantitative analysis of CT scans might plausibly be more objective than ‘qualitative’ analysis (i.e. short-ordered atrophy rating scales, with atrophy rated as none, mild, moderate or severe), where inter-observer reliability is only fair, even amongst expert neuroradiologists [17].

If clinicians could identify those people most likely to develop delirium based on clinical characteristics and neuroimaging on admission, efforts to prevent delirium could be focused on these patients. If trialists could identify those most at risk of delirium, this could help stratify delirium disk in clinical trials.

Thus, our aims were to explore the relationship between delirium after stroke and brain atrophy (using both qualitative atrophy assessments and quantitative linear measures), WMD, acute stroke lesion and previous stroke lesions on routinely performed CT brain scans, corrected for clinical covariables.

## Methods

### Design

This study was nested within a longitudinal cohort study of delirium after stroke in 95 patients admitted to an acute stroke unit in a teaching hospital from October 2012 for one year, and followed up for a year. Power calculations were based on the primary aim of determining associations between delirium and cortisol dysregulation; main results have already been reported [18]. Inclusion criteria were clinically confirmed stroke, age ≥ 60 years and onset within previous 120 hours (or time since last seen well for strokes occurring overnight) at the time of consent. Exclusion criteria were transient ischaemic attack, subarachnoid haemorrhage, current or recent use (within 6 months) of oral or inhaled corticosteroids, active alcohol withdrawal and inability to speak English. Patient or proxy consent was obtained by a clinical research fellow (AJB).

### Baseline clinical data

This included pathological stroke type, subtype (Oxfordshire community Stroke Project Classification) and stroke severity. Premorbid cognition was assessed using The Informant Questionnaire on Cognitive Decline in the Elderly (IQCODE) [19] and The National Adult Reading Test (NART) [20]. Heart rate, blood pressure, oxygen saturations and temperature, medications and admission blood results were recorded at the time of recruitment. The APACHE II (The Acute Physiology and Chronic Health Evaluation System II) was used as a measure of current illness burden [21].

### Delirium assessments

Participants were assessed for delirium by AJB at 1, 3, 5, 7, 14, 21, 28 days, 4 months and 12 months after stroke onset. Participants discharged after a short admission were seen as per protocol whilst an in-patient and were seen at home on day 28, at 4 months and 12 months. Delirium diagnosis was based on the four DSM-IV criteria (acute onset or fluctuating course, inattention, disorganised thinking and altered level of consciousness).

All participants underwent CT brain scanning as part of their routine clinical care using a Toshiba 64-slice or later in the study a 128-slice scanner. Scans were stored in the Patient Archiving and Communication System (PACS).

AJF (consultant neuroradiologist) used a standard method of extracting information on stroke lesion location, anterior and posterior white matter lesions using the Van Swieten scale [22] and global cerebral atrophy [23].

AJB was trained in quantitative brain imaging analysis by a Research Fellow (KF) with extensive experience of this methodology: linear measurements were taken directly from electronic images blind to all other participant data. Images were aligned to the AC-PC line and adjusted for symmetry. The graphics function of PACS was used for measurements [16] (figure 1). Measurements A, B, C, H, I, O, P and N were taken from the slice where the feature is most clearly visualised, measurement E, F and J were taken on the slice that shows most prominently the heads of the caudate nuclei, G is taken on the slice displaying most of the body of the lateral ventricle and M was taken from the top slices without displaying the lateral ventricle. Measurements took approximately 20 minutes for each scan. With the exception of measurements relating to skull size (A, B and C), a larger measurement indicated more atrophy.

**Figure 1.**
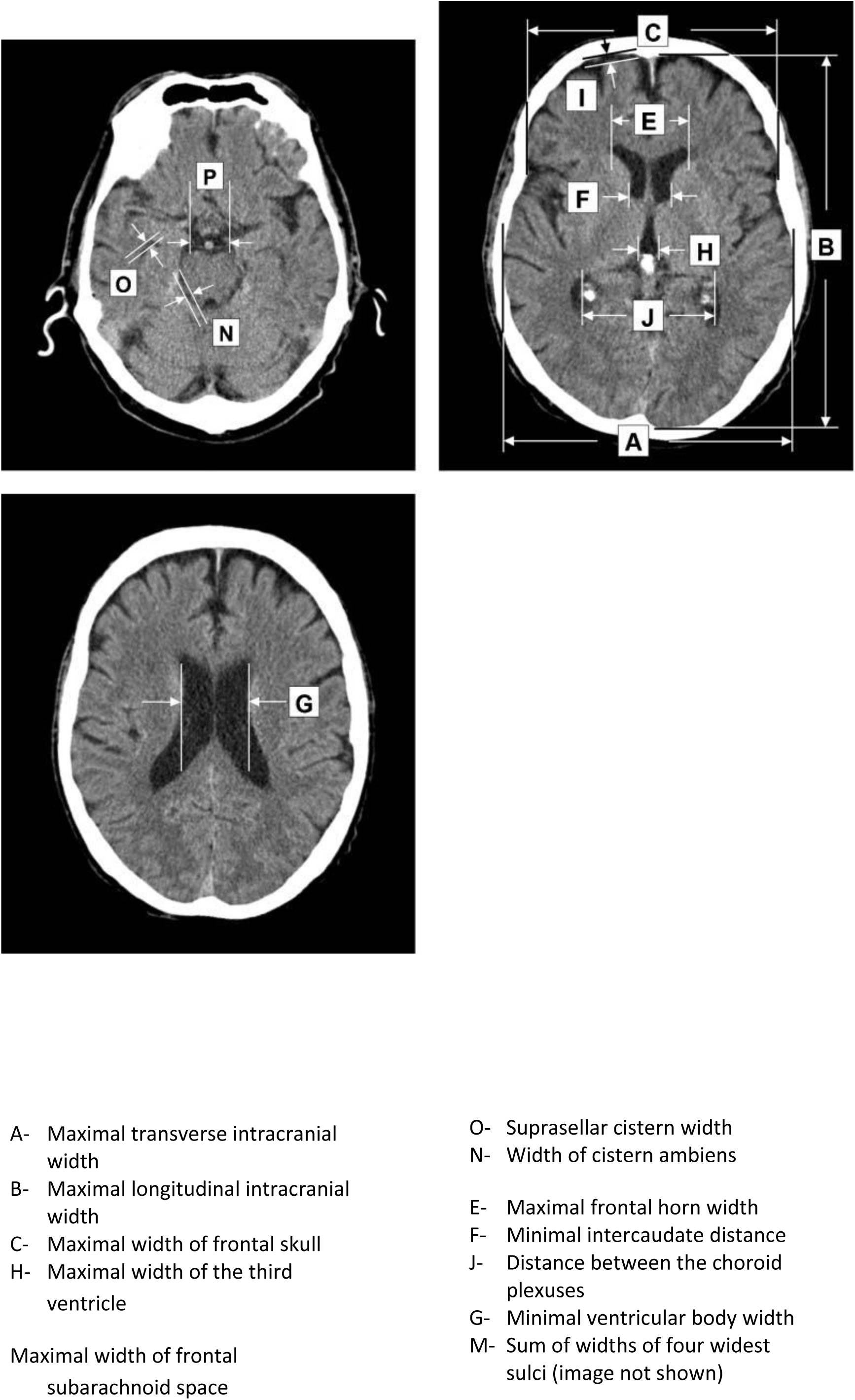
Linear measurements taken on axial CT slices Reproduced with permission from Acta radiologica (SAGE publications)

From these measurements, the following indices and ratios were calculated: Evans ratio (E/C), bicaudate ratio (F/A), Huckman number (E+F), cella media index (G/A), third ventricular ratio (H/A), ventricle index (J/E), frontal subarachnoid ratio (I/B), four cortical sulcal ratio (M/A), cistern ambiens ratio (N/A) temporal horn ratio (O/A) and supracellar cistern ratio (P/A). Those ratios which involve measurements A, B or C (measurements of the skull vault) are included as they correct for skull size (an indirect measure of peak brain size).

Intra-rater reliability was assessed by AJB repeating measurements from 10 participants 4 weeks later, blind to the results of the first assessment. Inter-rater reliability was tested for 20 participants by KF blind to AJB’s measurements.

### Statistical analysis

Non-parametric analysis was used because of short ordinal scales and the non-normal distribution of some data. The Mann-Whitney U test (for continuous data) or the chi-square test (for nominal categorical data) was used to investigate the relationship between delirium diagnosis and baseline characteristics.

For qualitative measurements, inter-rater and intra-rater reliability were tested by weighted kappa.

For linear measurements intra-reliability and inter-reliability were assesses using intraclass correlation (ICC) and Spearman’s correlations.

The relationship between a delirium diagnosis during the 12 month follow-up and qualitative CT assessments (presence of an acute ischaemic lesion, acute haemorrhage, hyperdense artery, global cerebral atrophy (none, mild, moderate or severe), WMLs (none, mild moderate or severe and presence of an old stroke lesion) was assessed using Pearson’s chi-square test. The independent contribution of each variable was assessed by binary logistic regression (enter method). Delirium (yes/no) was the dependent variable. Predictor variables selected *a priori*, were WMLs (linear scale where 0 = none, 1 = mild 2=moderate and 3=severe), atrophy (scale 0-4), presence of an acute stroke lesion and presence of an old stroke lesion. Co-variates selected *a priori* were age, sex, NIHSS score at admission (stroke severity) and IQCODE.

The relationship between delirium and linear CT measurements was initially investigated using point biserial correlations. All assumptions for multivariable logistic regression were tested and met. Then multivariable analysis was performed (enter method), with age, sex, NIHSS, IQCODE (variables selected *a priori*) and the linear measurements which had shown a significant correlation with delirium (p value <0.01) in the univariate analysis.

## Results

### Participants

All 95 participants had a CT brain scan; there are no missing data from these scans. Demographic data have already been reported [18]. Seventy-four participants were assessed at 4 months and 68 at 12 months (attrition was mainly due to death, ongoing ill health and disability). Twenty-six (27%) had delirium at some point during the 12 month follow-up. Delirium was diagnosed at a median of day 5 after stroke (IQR day 4-day 7) and median delirium severity score (DRS-R98) was 22 (IQR 16-26). Delirium was associated with increasing age, being female, less independent in pre-stroke ADLs, worse IQCODE, higher NIHSS and having had a TACS on univariable analysis (table 1).

**Table 1.**
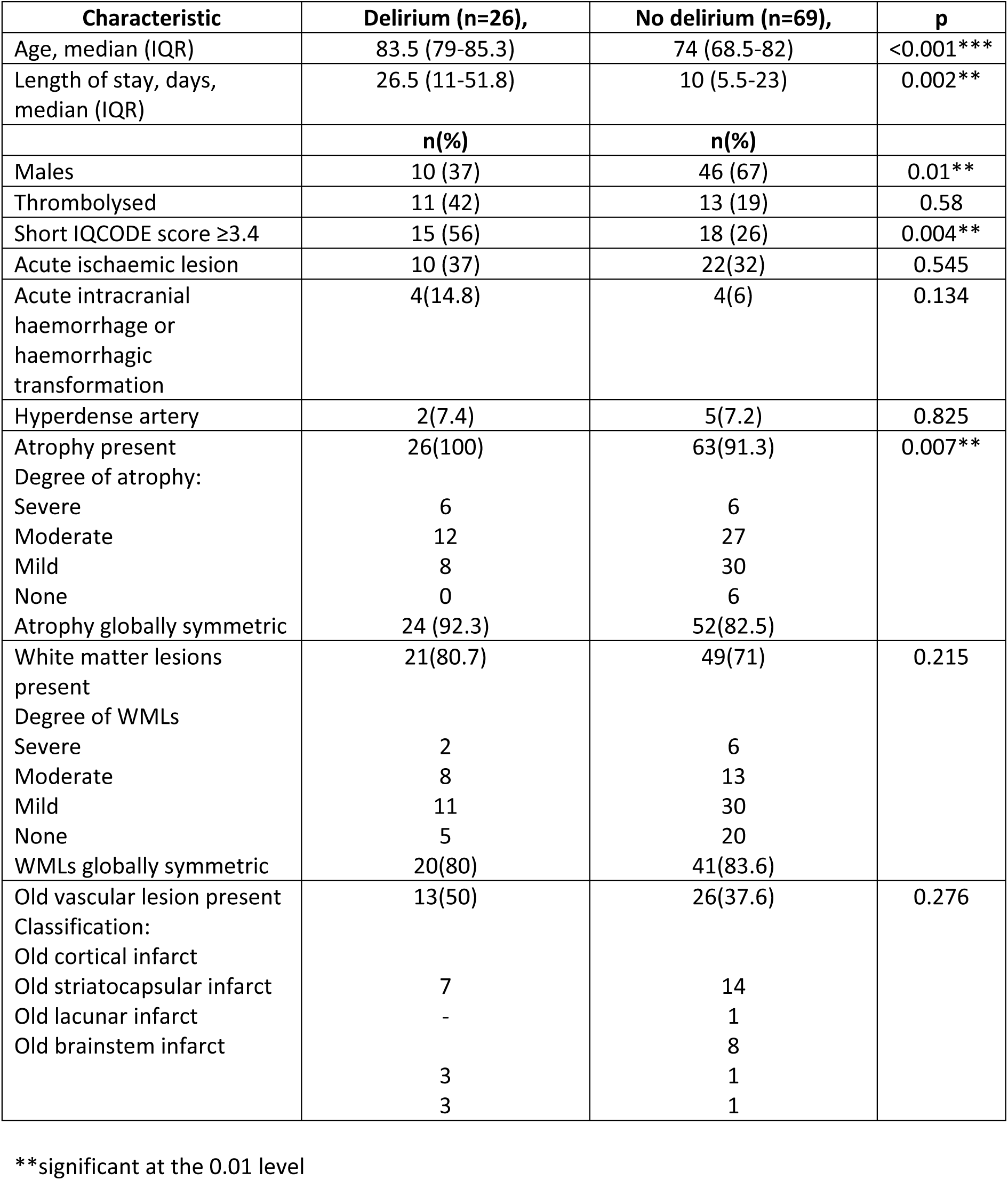
Descriptive statistics of participants with and without delirium in the 12 months after stroke.

| Characteristic | Delirium (n=26), | No delirium (n=69), | p |
| --- | --- | --- | --- |
| Age, median (IQR) | 83.5 (79-85.3) | 74 (68.5-82) | <0.001*** |
| Length of stay, days, median (IQR) | 26.5 (11-51.8) | 10 (5.5-23) | 0.002** |
|  | **n(%)** | **n(%)** |  |
| Males | 10 (37) | 46 (67) | 0.01** |
| Thrombolysed | 11 (42) | 13 (19) | 0.58 |
| Short IQCODE score $\geq 3.4$ | 15 (56) | 18 (26) | 0.004** |
| Acute ischaemic lesion | 10 (37) | 22(32) | 0.545 |
| Acute intracranial haemorrhage or haemorrhagic transformation | 4(14.8) | 4(6) | 0.134 |
| Hyperdense artery | 2(7.4) | 5(7.2) | 0.825 |
| Atrophy present | 26(100) | 63(91.3) | 0.007** |
| Degree of atrophy: |  |  |  |
| Severe | 6 | 6 |  |
| Moderate | 12 | 27 |  |
| Mild | 8 | 30 |  |
| None | 0 | 6 |  |
| Atrophy globally symmetric | 24 (92.3) | 52(82.5) |  |
| White matter lesions present | 21(80.7) | 49(71) | 0.215 |
| Degree of WMLs |  |  |  |
| Severe | 2 | 6 |  |
| Moderate | 8 | 13 |  |
| Mild | 11 | 30 |  |
| None | 5 | 20 |  |
| WMLs globally symmetric | 20(80) | 41(83.6) |  |
| Old vascular lesion present | 13(50) | 26(37.6) | 0.276 |
| Classification: |  |  |  |
| Old cortical infarct |  |  |  |
| Old striatocapsular infarct | 7 | 14 |  |
| Old lacunar infarct | - | 1 |  |
| Old brainstem infarct |  | 8 |  |
|  | 3 | 1 |  |
|  | 3 | 1 |  |

\*\*significant at the 0.01 level

### Qualitative assessment of the CT scans

An acute stroke lesion was visible in 40 (42%) participants (32 had ischaemic lesions and 8 had primary intracerebral haemorrhage (ICH)). Old vascular lesions were seen in 39 (41%), cerebral atrophy (mild, moderate or severe) in 89 (93.6%), and WMLs in 70 (73.6%). The atrophy and WMLs were symmetric in 85% of participants. One scan was entirely normal. On univariate analysis, delirium was associated with global cerebral atrophy (100% of those with delirium vs 91% without) (table 1). There was no relationship between the delirium and visible acute or chronic, ischaemic or haemorrhagic lesion, hyperdense artery or WMLs.

The multivariable model accounted for between 38% and 55% of the variance in delirium status (table 2). Being female, higher NIHSS score and the presence of atrophy were the only independent associations with delirium.

**Table 2.** Association between delirium and predictor variables (including qualitative brain CT) on multivariate analysis using binary logistic regression model.

| Variable | Odds ratio | 95% CI | p |
| --- | --- | --- | --- |
| Age (years) | 1.07 | 0.98-1.18 | 0.114 |
| Sex (female) | 6.99 | 1.44-33.89 | **0.016\*** |
| NIHSS | 1.28 | 1.09-1.51 | **0.003\*\*** |
| IQCODE score | 6.23 | 0.36-107.84 | 0.208 |
| Brain atrophy (0-3) | 3.70 | 1.15-11.88 | **0.028\*** |
| White matter lesions (0-3) | 0.73 | 0.33-1.39 | 0.427 |
| Visible acute stroke lesion | 2.36 | 0.62-8.93 | 0.207 |
| Visible old stroke lesion | 0.494 | 0.13-1.90 | 0.304 |

\*significant at the 0.05 level \*\*significant at the 0.01 level \*\*\*significant at the <0.001 level

The quantitative linear measurements were similar to those reported by Zhang and colleagues (Zhang, 2008) (table 3). Intra-rater agreement for quantitative measurements was mostly excellent except for the maximal width of frontal skull, C (fair to good agreement), width of the cistern ambiens, N (fair to good agreement) and the temporal horn diameter, O (poor agreement). (table 1 appendix). Inter-rater reliability (between AJB and KF) was excellent for maximal skull width and frontal skull width, minimal intercaudate distance, maximal width of the third ventricle and the sum of the widths of the four widest sulci. There was fair to good agreement for the maximal frontal horn width, minimal ventricular body width, minimal width of the frontal subarachnoid space and the temporal horn diameter. The remainder of the measurements showed poor interrater reliability (table 2 appendix).

**Table 3.** Linear measurements on CT brain scans in all participants.

| Linear measurement | Alphabetical annotation | Median (mm) | IQR (mm) |
| --- | --- | --- | --- |
| Maximal transverse intracranial width | A | 138.3 | 134.1-141.6 |
| Maximal longitudinal intracranial width | B | 157.1 | 151.4-161.6 |
| Maximal width of frontal skull | C | 106.2 | 101.5-111.9 |
| Maximal frontal horn width | E | 38.6 | 35.2-41.5 |
| Minimal intercaudate distance | F | 19.4 | 16.9-23.7 |
| Minimal ventricular body width | G | 27.0 | 23.8-31.9 |
| Maximal width of the third ventricle | H | 8.1 | 7.2-10.0 |
| Maximal width of frontal subarachnoid space | I | 4.0 | 3.5-5.1 |
| Distance between the choroid plexuses | J | 53.6 | 47.7-58.9 |
| Sum of the widths of the four widest sulci | M | 20.2 | 17.8-23.3 |
| Width of the cistern ambiens | N | 4.3 | 3.9-5.2 |
| Temporal horn diameter | O | 4.4 | 3.7-5.4 |
| Supracellar cistern width | P | 25.8 | 24.2-28.1 |
| Evans ratio | E/C | 0.4 | 0.3-0.4 |
| Bicaudate ratio | F/A | 0.14 | 0.12-0.17 |
| Huckman number | E+F | 59.3 | 52.9-64.9 |
| Cella media index | G/A | 0.2 | 0.17-0.23 |
| Third ventricular ratio | H/A | 0.06 | 0.05-0.07 |
| Ventricle index | J/E | 1.4 | 1.2-1.5 |
| Frontal subarachnoid ratio | I/B | 0.03 | 0.02-0.03 |
| Four cortical sulci ratio | M/A | 0.15 | 0.13-0.17 |
| Cistern ambiens ratio | N/A | 0.03 | 0.03-0.04 |
| Temporal horn ratio | O/A | 0.03 | 0.03-0.04 |
| Supracellar cistern ratio | P/A | 0.19 | 0.17-0.20 |

### Relationship between delirium and the linear measurements of atrophy in specific brain areas

We specified *a priori* that a p value of ≤0.01 would be considered significant. Thus, only the maximal longitudinal intracranial width and the cisterns ambiens ratio correlated significantly with delirium on univariable analysis (table 4). A logistic regression analysis was performed (table 5) with delirium as the dependant variable. Independent variables were age, sex, NIHSS, IQCODE and the linear measurements which had significantly correlated with delirium in the univariate analysis (p value <0.01). Only NIHSS score and the cistern ambiens ratio (N/A) were associated with delirium (95 cases analysed; Chi-square p < 0.0005). The model accounted for between 35.1% and 51.2% of delirium variance.

**Table 4.** Univariate analysis of relationship between delirium (yes/no) and linear measurements on CT brain.

| Linear measurement | Alphabetical annotation | $r_{pb}$ | p |
| --- | --- | --- | --- |
| Maximal transverse intracranial width | A | -0.157 | 0.128 |
| Maximal longitudinal intracranial width | B | -0.300 | **0.003\*\*** |
| Maximal width of frontal skull | C | -0.131 | 0.206 |
| Maximal frontal horn width | E | 0.132 | 0.204 |
| Minimal intercaudate distance | F | 0.113 | 0.278 |
| Minimal ventricular body width | G | -0.017 | 0.872 |
| Maximal width of the third ventricle | H | 0.197 | 0.057 |
| Maximal width of frontal subarachnoid space | I | 0.229 | 0.026 |
| Distance between the choroid plexuses | J | -0.113 | 0.277 |
| Sum of the widths of the four widest sulci | M | 0.101 | 0.328 |
| Width of the cistern ambiens | N | 0.208 | 0.043 |
| Temporal horn diameter | O | 0.181 | 0.080 |
| Supracellar cistern width | P | -0.017 | 0.869 |
| Evans ratio | E/C | 0.214 | 0.038 |
| Bicaudate ratio | F/A | 0.146 | 0.159 |
| Huckman number | E+F | 0.031 | 0.765 |
| Cella media index | G/A | -0.016 | 0.877 |
| Third ventricular ratio | H/A | 0.203 | 0.049 |
| Ventricle index | J/E | -0.228 | 0.027 |
| Frontal subarachnoid ratio | I/B | 0.240 | 0.020 |
| Four cortical sulci ratio | M/A | 0.135 | 0.193 |
| Cistern ambiens ratio | N/A | 0.323 | **0.001\*\*** |
| Temporal horn ratio | O/A | 0.190 | 0.067 |
| Supracellar cistern ratio | P/A | 0.013 | 0.898 |

$r_{pb}$ = point-biserial correlation coefficient. \*\*significant at the 0.01 level

**Table 5.** Multivariate analysis: Binary logistic regression model. Predictors of delirium at any time point after stroke (n=95)

| Predictor variable | Odds Ratio | CI | p |
| --- | --- | --- | --- |
| Maximal longitudinal intracranial width (B) | 0.90 | 0.80-1.00 | 0.190 |
| Cistern ambiens ratio (N/A) | 1.41 | 1.48-4.96 | **0.028\*** |
| Age (years) | 1.09 | 1.00-1.19 | 0.075 |
| Sex (being female) | 0.65 | 0.14-3.13 | 0.536 |
| IQCODE (positive or negative) | 1.83 | 0.13-25.74 | 0.653 |
| NIHSS score (per point) | 1.23 | 1.06-1.43 | **0.006\*\*** |

\*significant at the 0.05 level \*\*significant at the 0.01 level \*\*\*significant at the <0.001 level

## Discussion

Delirium up to 12 months after stroke was associated with brain atrophy but not WLM, and the presence of an acute or chronic stroke lesion on qualitative readings of brain CT. With each one point increase in atrophy score, participants were 3.36 times more likely to develop delirium. For quantitative linear atrophy measurements of specific brain areas, delirium was associated only with cistern ambiens ratio and NIHSS.

The finding that brain atrophy, but not WMLs, are associated with delirium after stroke is in keeping with findings of at least one study [8]. In another study, the extent and rate of total brain atrophy and ventricular enlargement were associated with development of dementia, independent of hippocampal atrophy [24], so it is possible that those with pre-existing brain atrophy are more vulnerable to developing delirium, even if they do not have dementia, perhaps by virtue of the neuroanatomical changes found in atrophic brains e.g. narrowed gyri, widened sulci and enlarged ventricles, which reflect neuronal loss [25]. Neuronal loss, perhaps in certain critical regions [26], may reduce the resilience of the brain to precipitants commonly implicated in delirium. However, the role of atrophy is not certain-as a more recent population-based study of stroke found that only WML, and not atrophy, predicted subsequent delirium [13].

We found no relationship between a visible stroke lesion and delirium; our study might have been underpowered or an ischaemic lesion might not have become visible because all scans were performed within 24 hours of stroke onset. A higher NIHSS score was associated with delirium, in keeping with previous studies [8,27]. In our study, participants were assessed more frequently and over a longer period than these previous studies, meaning that prolonged episodes of delirium or incident delirium occurring after the first few days of stroke were unlikely to have been missed. In our multivariable analysis for both the qualitative assessment of atrophy and the quantitative linear measurements, delirium was not associated with age. In general, brain atrophy is associated with advancing age [28], however covariance testing did not find a significant relationship which would have precluded both age and atrophy being included in the model. Perhaps other factors e.g. pre-existing dementia, were more important in determining which participants had brain atrophy rather than age alone. However, several other well conducted studies which did not include brain atrophy in their analysis found associations between age and delirium after stroke [27,6, 29].

The finding that the cistern ambiens ratio is the only quantitative measurement that predicts delirium is intriguing. However, there was poor inter-rater agreement for the cistern ambiens ratio, in multivariate analysis, the 95% confidence intervals were wide, and there was only a small change in cistern ambiens size per unit change in delirium status. Possible reasons include small distances leading to poorer accuracy; reorienting the scan to ensure symmetry introduces error on re-measurement; anatomical boundaries not well defined and there were small numbers of scans for reliability. Zhang and colleagues found an association between Alzheimer’s disease and a greater temporal horn ratio and supracellar cistern ratio compared to controls [16]; this is in keeping with medial temporal lobe atrophy, a pathological change in Alzheimer’s disease. The ambient cistern is part of the subarachnoid cisterns, and widening of the cistern results from atrophy of the structures which border it [30]. Furthermore, the cistern ambiens laterally extends to become the transverse fissure, which is one of the perihippocampal fissures [30]. Atrophy of the perihippocampal fissures is associated with ageing and Alzheimer’s Disease [31].

It is not the absolute measurement of the cistern ambiens that was associated with delirium in this study, but rather the ratio of the cistern to the maximum transverse intracranial width. Head size is fixed throughout adulthood. Intracranial area varies considerably between individuals and is an estimate of peak or maximal brain volume [32], which is attained at around 6 years of age [33]. This is important as whole brain volume correlates positively with current intelligence [32] and intracranial area correlates positively with general cognitive ability [33]. Thus the associations we found with delirium may simply reflect a participant’s cognitive reserve, as a function of their general cognitive ability.

The other linear measurements were not associated with delirium, even though the qualitative atrophy rating was. Quantification produces a number which can be difficult to interpret unless there are also longitudinal measurements, although measuring CSF spaces is less ambiguous than regional brain volumes. Qualitative ratings indicate change from baseline so may better represent pathological changes.

This study has several strengths: firstly, our delirium assessments performed by the clinical research fellow were methodical, detailed, well validated, included subjective and objectives methods of diagnosis, and DSM IV criteria were used. Assessments were carried out sufficiently regularly that it’s unlikely an episode of delirium was missed. Particular attention was paid to the features of hypoactive delirium (for example reduced level of arousal) which is easy to overlook. Furthermore, delirium assessments were also performed at follow-up visits and, where possible, episodes of delirium between assessment points at 4 months and 12 months were ascertained from informants, clinical notes and general practitioners.

Secondly, patients with impaired conscious levels were included; this is essential, because drowsiness is a common feature of delirium. Delirium diagnosis relies on identifying disorganized thinking and impaired attention; by performing very detailed delirium assessments we were able to apply DSM-IV criteria to all participants including those with aphasia.

Thirdly, using data from routinely collected CT brain scans meant that we could include scan data from all participants, enhancing the external validity of the study. Finally, the median NIHSS score of 5 (range 1-28, IQR 4-8) was clinically representative of stroke severity in the UK at the time of the study.

There are some limitations: the study was relatively small (95 participants), however, in the field of neuroimaging in delirium research, this is still an important addition to the literature. Recruiting patients into this type of study requires researchers to approach frail, elderly, unwell patients without mental capacity to consent. There are no benefits in terms of receiving a potential new treatment, but detailed follow-up may be perceived to be beneficial particularly after hospital discharge.

Only 5% of participants had an ICH. There are several barriers to recruiting people with an ICH including a four-fold increase in the risk of death compared to those with ischaemic stroke and higher stroke severity. We controlled for undiagnosed dementia using IQCODE [19], but some pre-existing undiagnosed dementia may have been missed. There was attrition of participants, particularly from the group who developed delirium. Thus, those with the most severe strokes might have been more likely to die than develop delirium.

There are several avenues for future research. As CT technology improves, scans will become more fine-grained, and this presents opportunities to map any acute structural changes, including any that occur during an episode of delirium, in crucial regions such as the hippocampus. A systematic review and meta-analysis of observational studies in delirium after stroke, which includes our study, would be of value. Data could be combined from studies with different designs using Harvest plots and explore the impact of correction (or not) for confounding variables. Given that delirium predicts poorer cognitive outcomes, that brain imaging findings predict poorer outcomes, and that brain imaging is associated with delirium-a key question is whether delirium directly causes cognitive impairment or whether it’s simply a marker of a vulnerable brain. This is important because this would lead to trials to treat the underlying brain disease, or to treat the delirium-to prevent longer term cognition. Large studies are needed for this to ensure sufficient power-routinely collected clinical data, if CT results are available-using data linkage would be one way to do this.

## Data Availability

Data may be available at reasonable request

## Acknowledgements

We are grateful to the participants and their families who kindly took part in the study, and the service user group who advised on study design. We are also grateful to the clinical staff who supported recruitment.

## Funding

Dr Amanda Barugh and this study was funded through a research training fellowship from the Dunhill Medical Trust, UK

## Disclosures

There are no relevant disclosures

## Appendices

**Table 1 appendix.** Intraclass correlation (ICC) and Spearman’s correlations for intrarater reliability for linear measurements.

| Linear Measurement | Alphabetical annotation | Spearman's rho | p | ICC |
| --- | --- | --- | --- | --- |
| Maximal transverse intracranial width | A | 0.823 | **0.003\*\*** | 0.931<sup>+++</sup> |
| Maximal longitudinal intracranial width | B | 0.927 | **<0.001\*\*\*** | 0.915<sup>+++</sup> |
| Maximal width of frontal skull | C | 0.539 | 0.108 | 0.685<sup>++</sup> |
| Maximal frontal horn width | E | 0.903 | **<0.001\*\*\*** | 0.881<sup>+++</sup> |
| Minimal intercaudate distance | F | 0.673 | **0.033\*** | 0.861<sup>+++</sup> |
| Minimal ventricular body width | G | 0.770 | **0.009\*\*** | 0.953<sup>+++</sup> |
| Maximal width of the third ventricle | H | 0.600 | 0.067 | 0.882<sup>+++</sup> |
| Maximal width of the frontal subarachnoid space | I | 0.636 | **0.048\*** | 0.755<sup>+++</sup> |
| Distance between the choroid plexuses | J | 0.915 | **<0.001\*\*\*** | 0.780<sup>+++</sup> |
| Sum of the widths of the four widest sulci | M | 0.661 | **0.038\*** | 0.828<sup>+++</sup> |
| Width of cistern ambiens | N | 0.467 | 0.174 | 0.660<sup>++</sup> |
| Temporal horn diameter | O | 0.273 | 0.446 | 0.227<sup>+</sup> |
| Supracellar cistern width | P | 0.758 | **0.011\*\*** | 0.771<sup>+++</sup> |

\*significant at the 0.05 level
\*\*significant at the 0.01 level
\*\*\*significant at the <0.001 level
<sup>+</sup>Poor agreement
<sup>++</sup>Fair to good agreement
<sup>+++</sup>Excellent agreement

**Table 2 appendix.**
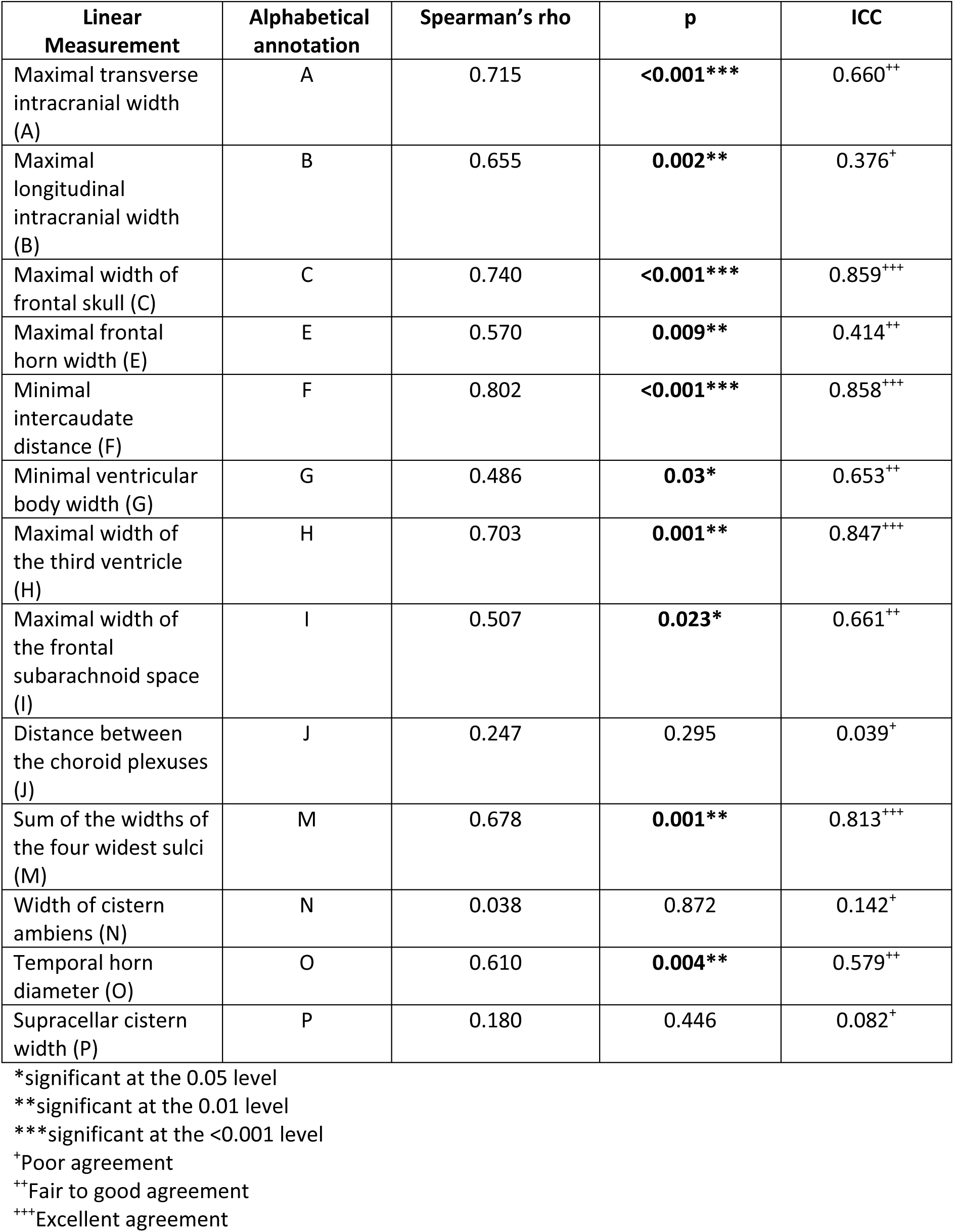
Intraclass correlation (ICC) and Spearman’s correlations for interrater reliability for linear measurements.

| Linear Measurement | Alphabetical annotation | Spearman’s rho | p | ICC |
| --- | --- | --- | --- | --- |
| Maximal transverse intracranial width (A) | A | 0.715 | <0.001*** | 0.660<sup>++</sup> |
| Maximal longitudinal intracranial width (B) | B | 0.655 | 0.002** | 0.376<sup>+</sup> |
| Maximal width of frontal skull (C) | C | 0.740 | <0.001*** | 0.859<sup>+++</sup> |
| Maximal frontal horn width (E) | E | 0.570 | 0.009** | 0.414<sup>++</sup> |
| Minimal intercaudate distance (F) | F | 0.802 | <0.001*** | 0.858<sup>+++</sup> |
| Minimal ventricular body width (G) | G | 0.486 | 0.03* | 0.653<sup>++</sup> |
| Maximal width of the third ventricle (H) | H | 0.703 | 0.001** | 0.847<sup>+++</sup> |
| Maximal width of the frontal subarachnoid space (I) | I | 0.507 | 0.023* | 0.661<sup>++</sup> |
| Distance between the choroid plexuses (J) | J | 0.247 | 0.295 | 0.039<sup>+</sup> |
| Sum of the widths of the four widest sulci (M) | M | 0.678 | 0.001** | 0.813<sup>+++</sup> |
| Width of cistern ambiens (N) | N | 0.038 | 0.872 | 0.142<sup>+</sup> |
| Temporal horn diameter (O) | O | 0.610 | 0.004** | 0.579<sup>++</sup> |
| Supracellar cistern width (P) | P | 0.180 | 0.446 | 0.082<sup>+</sup> |

\*significant at the 0.05 level \*\*significant at the 0.01 level \*\*\*significant at the <0.001 level <sup>+</sup>Poor agreement <sup>++</sup>Fair to good agreement <sup>+++</sup>Excellent agreement

## Notes

### Competing Interest Statement

The authors have declared no competing interest.

### Author Declarations

Scotland A Research ethics committee

